# AI-based image quality assessment in CT

**DOI:** 10.1101/2022.07.04.22277205

**Authors:** Lars Edenbrandt, Elin Trägårdh, Johannes Ulén

**Affiliations:** Region Västra Götaland, Sahlgrenska University Hospital, Department of Clinical Physiology, Gothenburg, Sweden; Department of Molecular and Clinical Medicine, Institute of Medicine, Sahlgrenska Academy, University of Gothenburg, Gothenburg, Sweden; SliceVault AB, Malmö, Sweden; Department of Clinical Physiology and Nuclear Medicine, Skåne University Hospital and Lund University, Malmö, Sweden; Wallenberg Center for Molecular Medicine, Lund University, Malmö, Sweden; Eigenvision AB, Malmö, Sweden

**Author notes:** Corresponding author Lars Edenbrandt.

**Keywords:** Artificial Intelligence, machine learning, quality assessment, diagnostic imaging

## Abstract

Medical imaging, especially computed tomography (CT), is becoming increasingly important in research studies and clinical trials and adequate image quality is essential for reliable results. The aim of this study was to develop an artificial intelligence (AI)-based method for quality assessment of CT studies, both regarding the parts of the body included (i.e. head, chest, abdomen, pelvis), and other image features (i.e. presence of hip prosthesis, intravenous contrast and oral contrast).

**Approach:** 1, 000 CT studies from eight different publicly available CT databases were retrospectively included. The full dataset was randomly divided into a training (*n* = 500), a validation/tuning (*n* = 250), and a testing set (*n* = 250). All studies were manually classified by an imaging specialist. A deep neural network network was then trained to directly classify the 7 different properties of the image.

**Results:** The classification results on the 250 test CT studies showed accuracy for the anatomical regions and presence of hip prosthesis in the interval 98.4% to 100.0%. The accuracy for intravenous contrast was 89.6% and for oral contrast 82.4%.

**Conclusions:** We have shown that it is feasible to develop an AI-based method to automatically perform a quality assessment regarding if correct body parts are included in CT scans, with a very high accuracy.

## 1. INTRODUCTION

Medical imaging, especially computed tomography (CT), is becoming increasingly important in research studies and clinical trials. Large projects and trials could include hundreds or thousands of CT studies and adequate image quality is essential for reliable results. This is a particular concern in multi-center trials, which often provide detailed imaging guides that must be followed in order to correctly include patients. Problems related to imaging could lead to either exclusion of patients or false image data incorporated in study or trial results. A quality check of images selected for a study is therefore an important process. Today, this is performed manually. Often, the quality check must be performed promptly by the clinical research organization before the patient can be enrolled. In both clinical trials and in large retrospective studies, this could be tedious work. The quality check of images could ensure for example that:

- the correct part of the body is visible in the CT, e.g., the chest in a lung cancer study,
- CT artefacts are not present, e.g., hip prostheses causing artefacts which usually prevent a proper analysis of the prostate,
- the CT is acquired according to the study protocol regarding the use of intravenous or oral contrast.

Information about a CT study should ideally be described in the DICOM tags. However, experience shows that it is not possible to rely only on this information. This is true especially in research projects and clinical trials when important information in the DICOM tags could be deleted in the pseudonymization process.

The use of artificial intelligence (AI) to solve clinical problems has been intensely studied in recent times.^1^ Deep learning, in particular, has gained attention as a method of obtaining complex information from medical images. AI could potentially be trained to help with image quality assessment and could be an important tool in this important, otherwise manual, task.

The aim of this study was to develop an AI-based method for quality assessment of CT studies, both regarding the parts of the body included (i.e. head, chest, abdomen, pelvis), and other image features (i.e. presence of hip prosthesis, intravenous contrast and oral contrast).

## 2. METHODS

### 2.1 Patients

We retrospectively included 1,000 CT studies from eight publicly available CT databases, see Table 1.

**Table 1.**
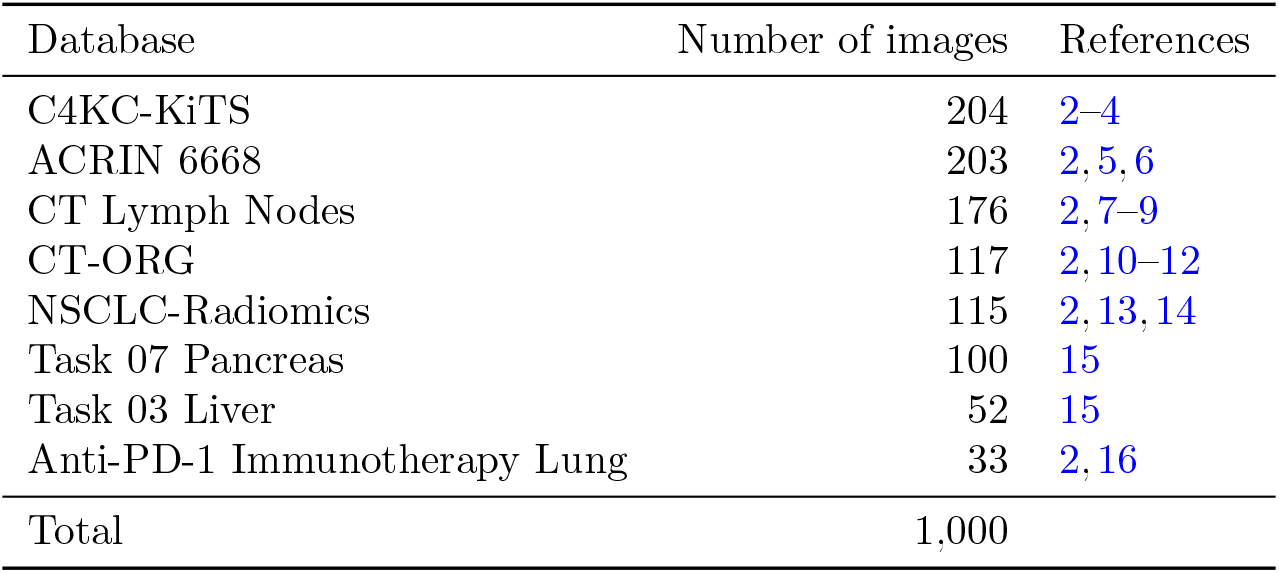
The number of images selected from each publicly available database.

| Database | Number of images | References |
| --- | --- | --- |
| C4KC-KiTS | 204 | 2–4 |
| ACRIN 6668 | 203 | 2, 5, 6 |
| CT Lymph Nodes | 176 | 2, 7–9 |
| CT-ORG | 117 | 2, 10–12 |
| NSCLC-Radiomics | 115 | 2, 13, 14 |
| Task 07 Pancreas | 100 | 15 |
| Task 03 Liver | 52 | 15 |
| Anti-PD-1 Immunotherapy Lung | 33 | 2, 16 |
| Total | 1,000 |  |

Before training the model, the full image set of 1, 000 CT studies was randomly divided into a training (*n* = 500), a validation/tuning (*n* = 250), and a testing set (*n* = 250). The test set was reserved for model evaluation.

### 2.2 Manual classification/Ground truth definition

All CT studies were classified by a nuclear medicine specialist experienced in hybrid imaging. Each case was classified based on the presence of the following seven features:

- *Head* The cranium is visible at least partly. Head is not present if only part of the mandible is visible.
- *Chest* The lungs are visible. Only very minor parts may be missing.
- *Abdomen* Main parts of liver, spleen, and the kidneys are visible.
- *Pelvis* The hip bones are visible.
- *Hip prosthesis* Uni- or bilateral hip prosthesis including implants for fixation of hip fractures.
- *Intravenous (IV) contrast* Signs of intravenous contrast including different phases.
- *Oral contrast* : Signs of oral contrast including different phases.

An overview of the distribution of the different classes in the dataset is given in Table 2.

**Table 2.** Positive and negative examples for each class in the dataset.

| Class | Positive count | Negative count |
| --- | --- | --- |
| Head | 256 | 744 |
| Chest | 603 | 397 |
| Abdomen | 903 | 97 |
| Pelvis | 702 | 298 |
| Hip prosthesis | 25 | 975 |
| Intravenous contrast | 256 | 744 |
| Oral contrast | 422 | 578 |

### 2.3 AI tool

The AI tool consists of a 3D-ResNet,^17^ a deep neural network designed for classification of 3D images. The network have an input shape of 110 × 110 × 110 × 1 pixels with 7 output channels each with its own sigmoid activation. Each output channel represents one of the classes defined in Section 2.2.

Many CT images contain quite a lot of air, which is not helpful for classification. In order to remove air, the images are smoothed using a Gaussian kernel with standard deviation 5mm^3^. An axis-aligned bounding box is then fitted to all pixels with Hounsfield unit (HU) above –800 in the smoothed image. The original image is then cropped to this bounding box.

The cropped CT images are pre-processed by clamping the HU values to the range [-1000, 3000] and then normalized to [-1, 1]. Furthermore, the CT volumes are re-sized to resolution 5 × 6 × 12 mm (or the smallest possible pixel shape with the same aspect ratio making the full image fit) and placed in the middle of the input volume.

#### 2.3.1 Sampling

The classes are quite imbalanced as seen in Table 2. In order to sample uncommon examples more often each image *i* is sampled proportional to a weight *w*_*i*_ defined as:

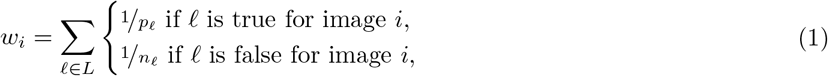

where *L* is the set of labels, *p*_*ℓ*_ the total number of positive examples, and *n*_*ℓ*_ the total number of negative examples for label *ℓ. p*_*ℓ*_ and *n*_*ℓ*_ are calculated individually for the training and validation sets.

#### 2.3.2 Training

Binary cross-entropy is used as loss function and the network is optimized using the ADAM optimizer^18^ with Nesterov momentum and an initial learning rate of 1 × 10^−5^. Each training and validation epoch consists of 2, 000 samples and 400 samples respectively. If the validation loss has not improved for 10 epochs the learning rate is halved until it reaches a minimum value of 1 × 10^−8^. The training stops when validation loss has not improved for 20 epochs.

During training the images are augmented using rotations (−0.1 to 0.1 radians), scaling (−10 to 10%) and an intensity shift of (−100 to +100 HU).

## 3. RESULTS

The classification results on the test set of is presented in Table 3. The accuracy for the anatomical regions and presence of hip prosthesis were 98.4% to 100.0%. The accuracy for intravenous contrast was 89.6% and for oral contrast 82.4%. Figure 1 and 2 show patient examples with correct and non-correct classifications.

**Table 3.** Result for the 250 test CT studies. True positive (TP), True negative (TN), False positive (FP), False negative (FN), and accuracy.

| Classification task | TP | TN | FP | FN | Accuracy |
| --- | --- | --- | --- | --- | --- |
| Head | 70 | 176 | 0 | 4 | 98.4% |
| Chest | 146 | 104 | 0 | 0 | 100.0% |
| Abdomen | 222 | 24 | 0 | 4 | 98.4% |
| Pelvis | 167 | 80 | 0 | 3 | 98.8% |
| Hip prosthesis | 6 | 244 | 0 | 0 | 100.0% |
| Intravenous contrast | 149 | 75 | 13 | 13 | 89.6% |
| Oral contrast | 71 | 135 | 15 | 29 | 82.4% |

**Figure 1.**
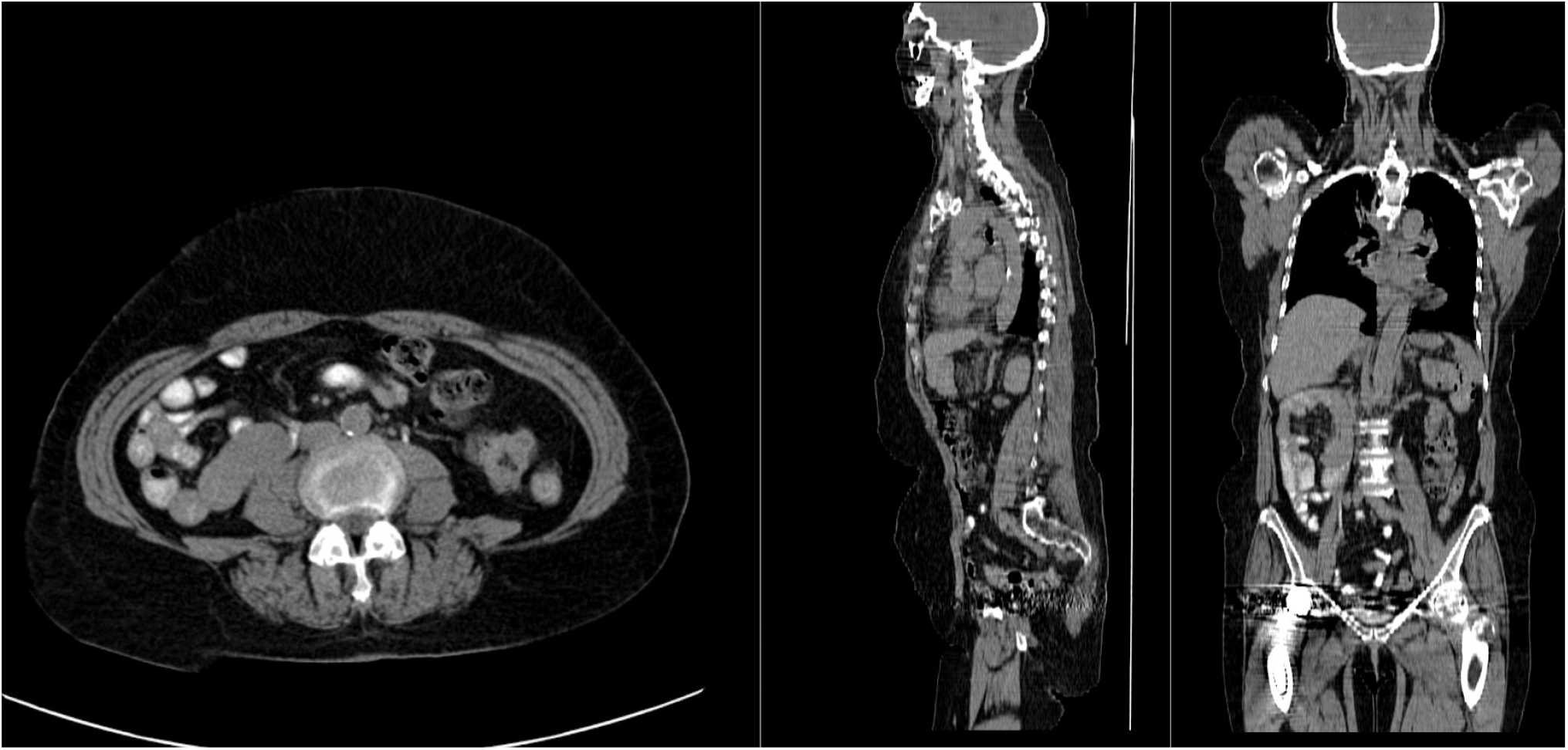
Example of a correctly classified image from ref. 5. Head, chest, abdomen, pelvis, hip prosthesis, and oral contrast were present. Intravenous contrast was not present.

**Figure 2.**
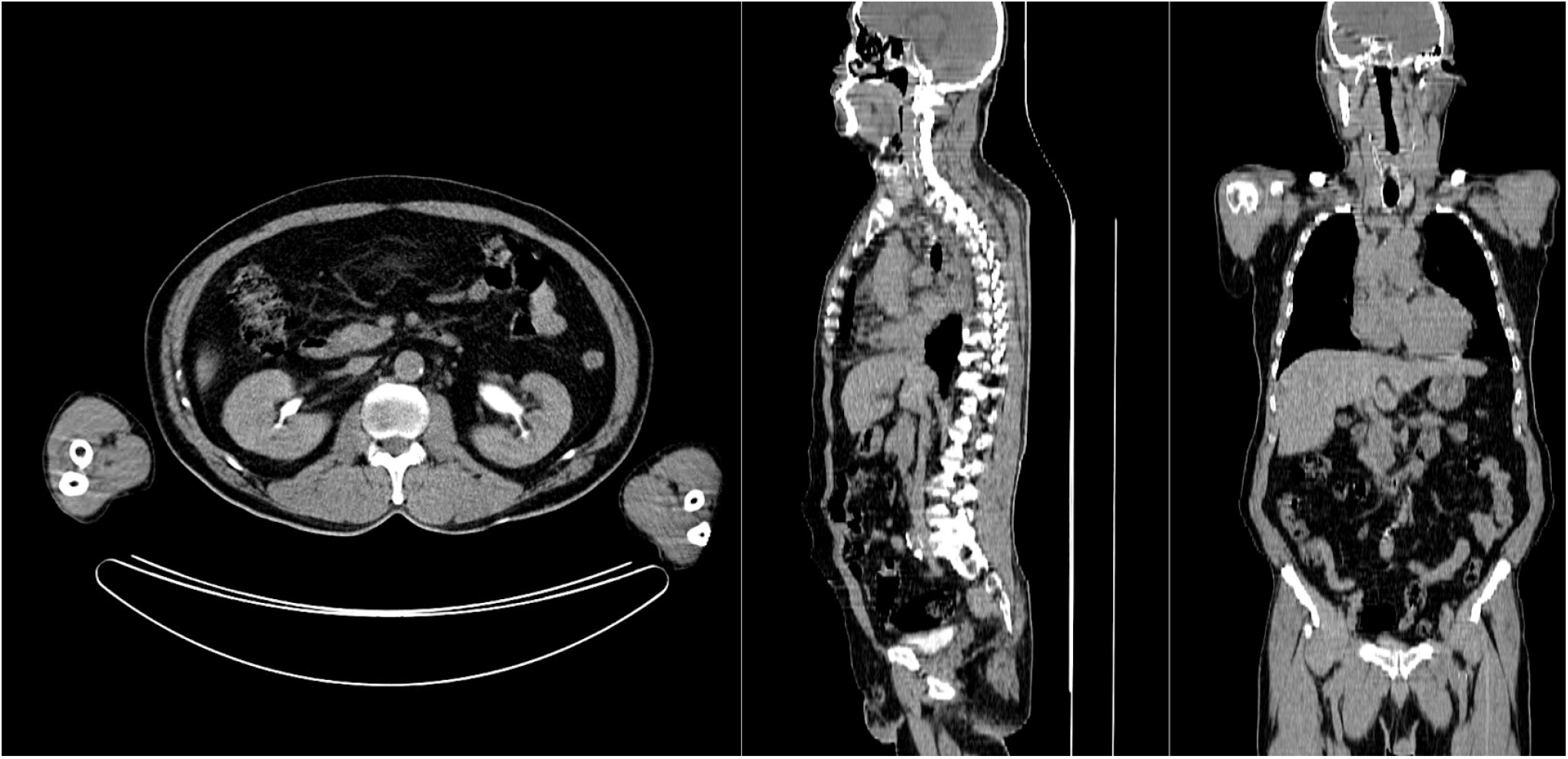
Example of image with a miss-classification from ref. 5. Excretory intravenous contrast phase is present, but not detected by AI. All other classifications were correct.

The execution time on a high-end desktop computer with a Nvidia RTX 3090 graphics card is about 2 seconds per image.

## 4. DISCUSSION

In this study we have shown that it is feasible to develop an AI-based tool to automatically check that the correct body parts are visible in the CT studies, with a very high accuracy. The AI-based method was also able to accurately detect hip prosthesis even though the number of positive cases in the training and validation sets were limited (*n* = 19).

A limitation of this study was that the AI-based tool only made a classification regarding presence of contrast or not. Many different phases of contrast enhancement exist,^19^ typically early arterial phase (15–25 s post injection), late arterial phase (30–40 s post injection), hepatic or late portal venous phase (70–90 s post injection), nephrogenic phase (85–120 s post injection) and excretory or delayed phase (5–10 min post injection). No clearly defined times post injection of the contrast agent exist, but with a large number of images with different contrast phases in the training group, it would probably be possible to train an AI-method to categorize the contrast phase in more detail than we did in this study. Other potential problems related to intravenous contrast is different amounts of contrast agent administered, for example reduced doses in patients with kidney disease. Problems with oral contrast for this type of task include different timings of contrast as well as different contrast agents (for example barium or iodine-based agents). A more comprehensive classification of contrast would most likely require a much larger training set. Some of the false negative cases of our test set represented very late intravenous phases with low contrast in the aorta but contrast in the kidneys or urinary bladder (Figure 2). This type of cases was not common in the training set. Also the appearance of oral contrast on CT showed substantial variation. In most cases contrast was clearly visible in the small bowel. In other cases only the stomach or colon showed contrast.

Medical imaging is often a key asset in clinical trials, as it can provide efficacy evaluation and safety monitoring.^20^ It is also often used as screening for eligible patients to include. Medical imaging can also improve clinical trial efficacy and reduce the time to complete a specific trial, by offering imaging biomarkers that can act as a surrogate endpoint. In order to do so, good image quality is crucial and therefore it is necessary to monitor image quality throughout different stages of a study. A step in the quality assessment could be to determine if correct body parts are included and if the images contain contrast agent or not. Further development of automated image quality assessment could also include image properties such as noise level and patient motion. Evaluation by a human observer is both time consuming and subjective. AI-based tools could help minimize both issues.

## 5. CONCLUSIONS

We have shown that it is feasible to develop an AI-based method to automatically perform a quality assessment regarding if correct body parts are included in CT scans, with a very high accuracy.

## Data Availability

All data produced in the present study are available upon reasonable request to the authors

https://www.cancerimagingarchive.net/

## 6. ACKNOWLEDGMENT

We would like to thank Måns Larsson and Olof Enqvist for fruitful discussions regarding this study and closely related topics.

